# Comparative Assessment of p16/Ki-67 Dual Staining Technology for Cervical Cancer Screening in Women Living with HIV (COMPASS-DUST) – Study Protocol

**DOI:** 10.1101/2022.11.10.22282170

**Authors:** Kehinde S. Okunade, Kabir B. Badmos, Austin Okoro, Yewande O. Ademuyiwa, Yusuf A. Oshodi, Adebola A. Adejimi, Nicholas A. Awolola, Oluchi Ozonu, Hameed Adelabu, Gbenga Olorunfemi, Alani S. Akanmu, Adekunbiola A. Banjo, Rose I. Anorlu, Jonathan S. Berek

**Affiliations:** Department of Obstetrics & Gynaecology, College of Medicine, University of Lagos, Lagos, Nigeria; Department of Obstetrics & Gynaecology, Lagos University Teaching Hospital, Lagos, Nigeria; Center for Clinical Trial, Research, and Implementation science (CCTRIS), College of Medicine, University of Lagos, Lagos, Nigeria; Department of Anatomic and Molecular Pathology, College of Medicine, University of Lagos, Lagos, Nigeria; Department of Nursing Science, College of Medicine, University of Lagos, Lagos, Nigeria; Department of Obstetrics & Gynaecology, Lagos State University College of Medicine, Lagos, Nigeria; Department of Community Health & Primary Care, College of Medicine, University of Lagos, Lagos, Nigeria; Division of Epidemiology and Biostatistics, School of Public Health, University of Witwatersrand, Johannesburg, South Africa; Department of Haematology & Blood Transfusion, College of Medicine, University of Lagos, Lagos, Nigeria; Department of Obstetrics & Gynaecology, Stanford University School of Medicine, Stanford, USA

**Author notes:** **Corresponding author: E-mail:** (KSO).

**Keywords:** Cervical cancer, Cervical intraepithelial neoplasia, DUST, Screening, WLHIV

## Abstract

The risk of progression of low-grade (CIN1) to high-grade cervical intraepithelial neoplasia (CIN2/3) is 3–5 times higher for women living with HIV (WLHIV) than for HIV-negative women. Evidence suggests that the current cervical cancer screening methods perform less effectively in WLHIV. Molecular-based p16/Ki-67 dual staining technology (DUST) is a safe and rapid assay that could be used to detect CIN2/3 with higher sensitivity and specificity. The study in this protocol will evaluate the performance of p16/Ki-67 dual staining technology (DUST) in cervical cancer screening among WLHIV. We will conduct an intra-participant comparative study (*Phase 1*) to enrol n=1,123 sexually active WLHIV aged 25–65 years at two accredited adult HIV treatment centres in Lagos, Nigeria to compare the performance of DUST to the currently used screening methods (Pap smear, hr-HPV DNA, or VIA testing) in detecting high-grade CIN and cancer (CIN2+). Subsequently, a prospective cohort study (*Phase 2*) will be conducted by enrolling all the WLHIV who are diagnosed as having low-grade CIN (CIN1) in *Phase 1* for a 6-monthly follow-up for 2 years to detect the persistence and progression of CIN1 to CIN2+. The findings of this study may provide evidence of the existence of a better performance screening method for the primary and triage detection of CIN2+ in WLHIV. It may also demonstrate that this high-performance test can improve the long-term predictive accuracy of screening by extending the intervals between evaluations and thus decrease the overall cost and increase screening uptake and follow-up compliance in WLHIV.

**Author Summary:** As there is evidence to suggest that the currently used screening methods for cervical cancer are less effective in women living with HIV (WLHIV), the proposed study in this protocol will evaluate the performance of a molecular-based p16/Ki-67 dual staining technology (DUST) in the primary and HPV-triage detection of CIN2+ in WLHIV. Using an intra-participant comparative study design (*Phase 1*), n=1,123 sexually active WLHIV aged 25–65 years will be enrolled at two accredited adult HIV treatment centres in Lagos, Nigeria to compare the performance of DUST to the currently used screening methods (Pap smear, hr-HPV DNA, or VIA testing) in detecting CIN2+. In *Phase 2*, a prospective cohort study design will be used to enrol all WLHIV who are diagnosed as having CIN1 in *Phase 1* for a 6-monthly follow-up for 2 years to detect persistent CIN1 and progression of CIN1 to CIN2/3. Overall, this very promising molecular-based technology (DUST) could reduce the long-term cumulative screening cost and procedure-related anxiety and fear, and subsequent improvement in cervical cancer screening uptake and follow-up compliance in WLHIV. We anticipate that these will have significant public health impacts as this innovative method will offer a unique paradigm shift for future control of cervical cancer within an integrated HIV treatment setting in LMICs.

## Introduction

Cervical cancer is a major public health problem and is the fourth leading cause of cancer death in women worldwide accounting for an estimated 604,000 new cases and 342,000 deaths in 2020 (1). In Nigeria, the annual number of cervical cancer cases is 14,089 and the number of deaths is 8,240 (2). Invasive cervical cancer (ICC) is regarded as an AIDS-defining illness (3). Sub-Saharan Africa is the region most affected by the global human immunodeficiency virus (HIV) epidemic with an estimated 25 million cases (4) accounting for about 80% of the global burden of ICC (1). Persistent infection with genital high-risk human papillomavirus (hr-HPV) is a necessary cause of ICC (5) and therefore, women living with HIV (WLHIV), who are disproportionately affected by HPV infection (6,7) are also more likely to have persistent HPV infections and a more rapid progression to ICC (7). Nigeria currently ranks second, behind South Africa, in the global burden of HIV (8). The increased coverage of antiretroviral treatment (ART) leading to the improved life expectancy of WLHIV together with the poor screening and follow-up have contributed significantly to the high prevalence and incidence of ICC in Nigeria and other low- and middle-income countries (LMICs) (9). Integrated cervical cancer screening is now used in most settings including Nigeria as an effective measure to reduce the morbidity and mortality due to ICC in WLHIV (10).

The World Health Organization (WHO) recommends HPV DNA-based test as the preferred method to detect cervical intraepithelial neoplasia (CIN), and this is to replace the widely used Papanicolaou (Pap) smear and visual inspection with acetic acid (VIA) (11). HPV DNA testing has been adopted in most developed countries as the primary method for cervical cancer screening and is now gaining acceptance in most low- and middle-income countries (LMIC) including Nigeria (12). While the sensitivity of HPV DNA testing is high in WLHIV, its specificity is limited (21.0–55.7%) with further reductions associated with younger age, advanced immunosuppression, and shorter duration of ART (13). Therefore, when HPV DNA testing is used as the main screening method, women are prone to having a false-positive diagnosis, which causes an increase in the number of unnecessary colposcopies (14). This could then lead to a waste of resources and increased procedure-related anxiety in patients due to the fear of cancer (13).

Evidence, therefore, suggests that the current cervical cancer screening methods may perform less effectively in WLHIV (13,15–17). Annual screening with an initial colposcopy where resources are available is recommended in WLHIV (18,19) in contrast with the 3- or 5-yearly screening intervals in HIV-uninfected women when using Pap smear or HPV DNA testing, respectively. This results in poor screening uptake and follow-up compliance in these women (20,21) attributed mostly to the out-of-pocket payment for screenings and the psychosocial effects of awaiting a potentially unpleasant result. Therefore, there is a need for an alternative screening strategy that can better detect CIN with long-term predictive accuracy and reduced frequency of screening follow-ups in WLHIV compared to the currently used screening methods.

Although the exact mechanism by which long-term HIV infection may promote cervical carcinogenesis is still largely unknown, the molecular evolution in the development of CIN and ICC may produce an accurate detection marker in WLHIV. The rare simultaneous expression of the tumour suppressor protein (p16) and the cellular marker of proliferation (Ki-67) in the same cervical epithelial cell implies the deregulation of the cell cycle induced by HPV (22) which can serve as a marker of cellular transformation (23) and the presence of CIN2/3 (24–26). DUST (p16/Ki-67 dual staining technology) could be a better cervical cancer screening method in WLHIV in resource-limited settings due to its better sensitivity (91.9–97.2%) and specificity (82.1–95.2%) in detecting CIN2/3 lesions when compared to the currently available screening tests such as Pap smear, HPV DNA and VIA testing (23–27). We, therefore, propose the study – “COMParative ASSessment of p16/Ki-67 DUal Staining Technology in HIV-infected women (COMPASS-DUST)” which is aimed to evaluate the performance of DUST in detecting CIN2/3 and ICC (CIN2+) as a primary and HPV-triage test in WLHIV. We will also assess the 2-year performance of DUST in predicting the persistence of and progression of CIN1 to CIN2+ in WLHIV.

## Materials and Methods

### Study design and setting

The “COMPASS-DUST” is a two-phase study that will be conducted in two accredited adult HIV treatment clinics in Lagos, Nigeria – The Lagos University Teaching Hospital (LUTH) in Idi-Araba and the Nigerian Institute of Medical Research (NIMR) in Yaba, Lagos, Nigeria. The two clinics provide comprehensive care for people living with HIV (PLHIV) in Lagos and its surrounding states in Southwest Nigeria. Together, these clinics attend to ∼19,000 PLHIV each month and offer integrated care to sexually active women with HIV infection, including reproductive health services such as routine cervical cancer screening with either Pap smear, HPV DNA testing or VIA depending on availability and affordability.

### Study population and recruitment criteria

In *Phase 1*, we will conduct an intra-participant comparative cross-sectional study involving sexually active women with well-controlled HIV who are otherwise healthy and aged 25–65 years (28) at the participating sites. Women with suspicious cervical lesions, those with ongoing pregnancy or within 6 weeks of childbirth, those with a previous history of hysterectomy, HPV DNA testing or Pap smear within the past year, previous HPV vaccination, or history of ICC or therapy for benign or malignant cervical lesions will be excluded from enrollment. A total of 1,123 WLHIV will be required to achieve 80% power at a target significance level of 5%. This is based on a comparative specificity of 96.5% for DUST and 93.2% for Pap smear (29) respectively for CIN2+ detection in the primary and HPV-triage of WLHIV with adjustment made for an estimated non-response or data recording error rate of 10%.

In *Phase 2*, a prospective cohort study will be conducted to involve WLHIV and diagnosed with CIN1 in *Phase 1*. Women with the diagnosis of CIN2+ at baseline assessment, lack of willingness to continue in the study, and withdrawal of consent at any time during the follow-up period will be excluded. We will enrol all WLHIV who are diagnosed with CIN1 for a 6-monthly follow-up for 2 years. As there are no previous studies that assessed the risk of CIN progression based on DUST status, the data generated will enable us to formulate hypotheses for testing in future studies.

### Study procedures and data collection

In *Phase 1*, potentially eligible participants will be enrolled by members of the study team using the consecutive sampling technique [Figure 1]. Study intent and procedures will be introduced, and informed consent is obtained before any study procedure. An electronic data collection form designed on the REDCap database will collect information on socio-demographic variables, HIV history (duration of diagnosis), ART history (types and duration of use), immune status (latest CD4+ cells count and viral load), and other relevant information aimed at evaluating the risk factors for cervical cancer (30). Information on participants’ HIV history, ART use, and immune status will be extracted from the patient’s electronic database at the adult HIV clinics. The women will then undergo pelvic examination and cervical liquid-based cytology (LBC) sample collection using a Cytobrush that is broken off and placed into a ThinPrep vial containing PreservCyt Solution (Hologic Inc, Marlborough, MA 01752, USA) for sequential analysis by the study pathologists for human papillomavirus (HPV) DNA testing, Papanicolaou (Pap) smear, and DUST. All the participants will subsequently undergo visual inspection with acetic acid (VIA) using 3–5% acetic acid by a study nurse as recommended by the WHO (31) and interpreted according to IARC (International Agency for Research on Cancer) criteria (32). Any dense opaque aceto-white lesion observed near the squamo-columnar junction will be reported as VIA positive (VIA+). Thereafter, a colposcopic evaluation (reference gold standard) will be performed during which 3–5 different images will be taken for documentation and subsequent post-evaluation review by the study clinical team for quality control. The clinical team members will take a turn on each of the three selected days of the week to perform the colposcopic examination and the photographic images of the findings will be jointly reviewed at the end of each week for concurrence and quality control. Lesions suspicious for CIN, as per usual care, will be biopsied and collected in a 10% buffered formalin-containing recipient for transportation and confirmation by histopathologic analysis at the Anatomic and Molecular Pathology (AMPATH) laboratory. The histology results will be interpreted as normal, CIN1, or CIN2+.

**Figure 1:**
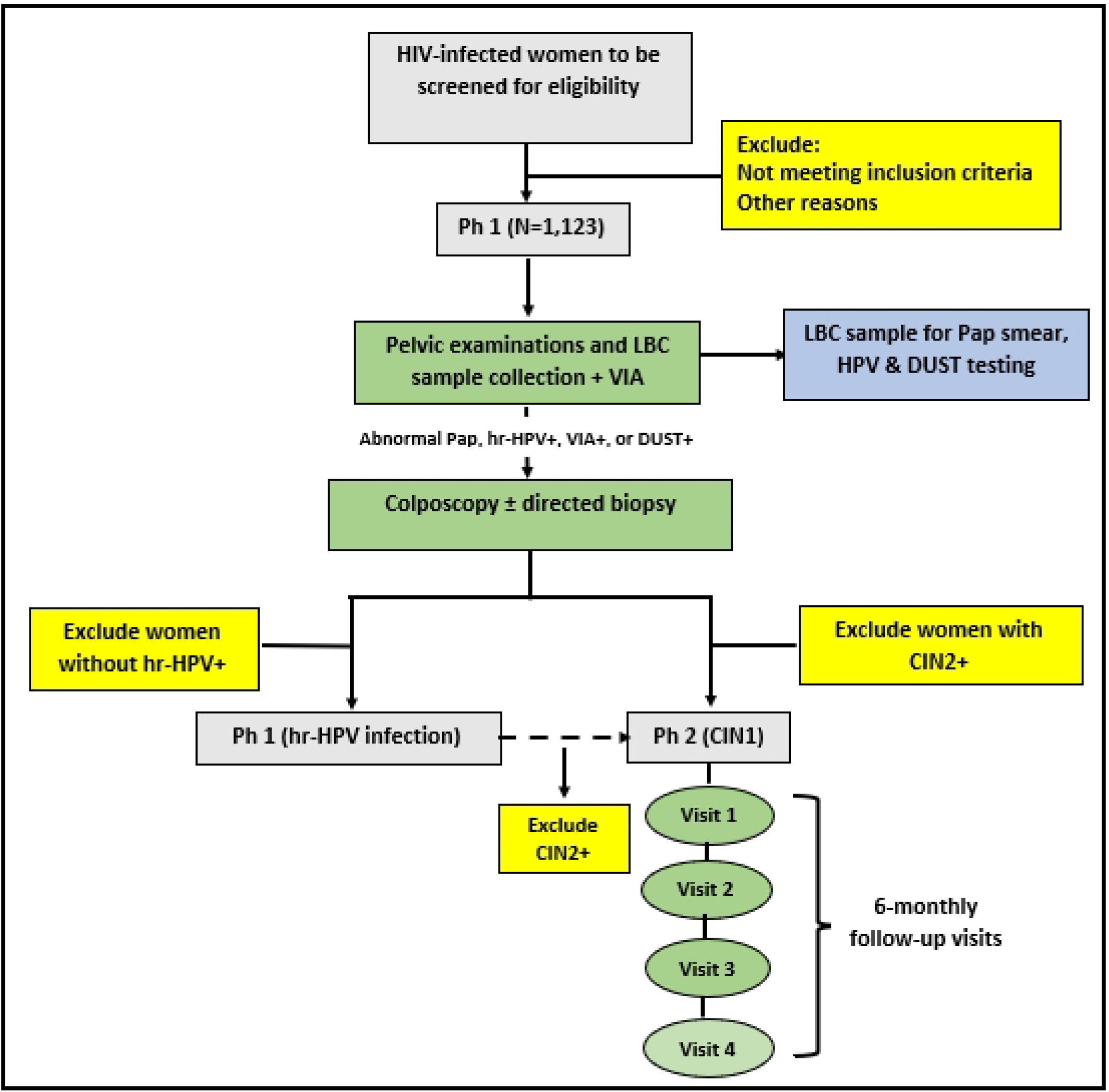
Study framework.

In *Phase 2*, all consecutively consenting WLHIV with the diagnosis of CIN 1 in *Phase 1*, will be identified and invited by the study investigators to participate. The enrolled women will undergo repeat Pap smear and hr-HPV DNA co-testing as per standard of care (33) at the 6th, 12th, 18th, and 24th months for a cumulative 2-year period. This is the minimum period required to see a difference in CIN progression between the screening methods (34). At enrolment, a participant’s locator form will be completed and then reviewed at each follow-up visit to ensure that the participant’s contact information is up to date. Women with ≥ASCUS or HPV+ or both will be recalled for further evaluation using colposcopy with(out) biopsy and histopathologic analysis. The study pathologists will be blinded to the baseline results or other follow-up data of the participants. Women with normal colposcopic (and/or histologic) findings will continue in the study while those with a positive test defined as CIN1 persisting till the last follow-up at the 24th month (persistent CIN1) or CIN2+ at any time will be offered standard treatment.

### Laboratory Analyses

Using the collected LBC sample, a study pathologist will perform the HPV testing using HPV DNA target amplification by Polymerase Chain Reaction (PCR) followed by nucleic acid hybridization for the detection of 14 high-risk HPV genotypes – HPV types 16, 18, 31, 33, 35, 39, 45, 51, 52, 56, 58, 59, 66 and 68. Two other study pathologists will subsequently read the other prepared slides from the PreservCyt residual LBC sample separately for Pap smear and DUST. The results of Pap smear cytology. will be classified according to the revised Bethesda’s classification (35) as negative for intraepithelial lesion and malignancy (NILM), Atypical squamous cells of undetermined significance (ASCUS), Atypical squamous cells cannot exclude HSIL (ASC-H), Low grade squamous intraepithelial lesion (LSIL) and High grade squamous intraepithelial lesion (HSIL). The slide for p16/Ki-67 dual staining (DUST) is prepared from the residual material using the ThinPrep 2000 Processor (Hologic) and will then be tested with the CINtec PLUS Cytology kit (manufactured by Roche mtm Laboratories AG, Mannheim, Germany). For the study, DUST tested sample is interpreted as either positive (DUST+) or negative (DUST-). Per the manufacturer’s instructions, CINtec PLUS detects a positive sample as the simultaneous appearance of brownish cytoplasmic staining (p16) and red nuclear staining (Ki-67) in one or more cells (DUST+), independent of cellular morphology. Slides without any double-stained cells will be labelled as negative for p16/Ki-67 dual staining (DUST-). A case will be scored as inadequate if adequate evaluation is impossible, no p16 and/or Ki-67 staining is visible as internal control, or if slides do not meet the squamous cellularity criteria as specified in the Hologic criteria (≥5000 cells per slide). Inadequate cases will be classified as negative (DUST-). The pathologist reading any slide will be blinded to the other screening tests or histology results, or other participants’ data.

### Statistical analysis

Data will be entered into the REDCap database and then exported to R Statistical Computing Software (R-4.1.2) for Windows for statistical analysis. Missing or incomplete data will be handled by running a missing values analysis using pairwise or listwise deletion if no pattern is detected or the multiple imputation (MI) method if a pattern is detected (36). Descriptive statistics will be used to describe the characteristics of the participating HIV-infected women and quantitative data will then be tested for normality with the Kolmogorov–Smirnov test with Lilliefors significance correction. Odds ratios and 95% confidence interval by 2×2 tables will be used to assess the association between each of the screening tests (DUST, Pap smear and HPV16/18) and histological outcomes. The sensitivity, specificity, positive predictive value, and negative predictive value of abnormal Pap smear (≥ASCUS), hr-HPV+, HPV16/18, or VIA+ as primary or HPV-triage tests will be compared to DUST. Multivariate analysis using a binary logistic regression model with the forward stepwise selection technique will be used to assess the associations between DUST+ and severity of cervical cytologic abnormalities and grade of CIN (CIN 1–3) while adjusting for other important covariates such as Age, ART history (type and duration), immune status (CD4+ cells count and viral load, etc.). Risks of persistence and progression of CIN1 will be compared with clinical management thresholds for colposcopy referral (and positive histologic diagnosis) and a 6-month return interval. Relative risks (RR) will be used to determine the strength of associations between participants’ baseline data and CIN1 progression (and persistence at 1- and 2 years). Women with <CIN1 at 2-year follow-up assessment will be censored. Longitudinal data will be evaluated using the multivariate Cox regression model with forward stepwise selection to test the association between baseline DUST status and progression of CIN1 to CIN2+ (and persistence of CIN1) at 1-year and 2-year. Adjustments will be made in the final multivariate models with the inclusion of other baseline variables with *P*<0.20 in the univariate analysis. Statistical significance for a two-tailed test will be reported at *P*<0.05.

## Discussion

This current study (COMPASS-DUST) will evaluate the diagnostic accuracy of a molecular-based marker (DUST) as a higher-performance primary and HPV-triage screening method for cervical cancer in WLHIV in comparison to the currently used screening methods such as Pap smear cytology, HPV DNA and VIA testing using the reference gold standard of colposcopy with(out) biopsy and histology for diagnostic confirmation. In addition, the study will also assess the 2-year performance of DUST in predicting the persistence of and progression of CIN1 to CIN2+ in WLHIV.

The findings of this study will be of great importance to public health practice for 2 reasons. First, they may provide evidence of the existence of a better performance screening method for the primary and triage detection of CIN2+ in WLHIV. Secondly, we may demonstrate that this high-performance test can improve the long-term predictive accuracy of screening by extending the intervals between evaluations. Overall, this very promising molecular-based technology (DUST) could reduce the long-term cumulative screening cost and procedure-related anxiety and fear, and subsequent improvement in cervical cancer screening uptake and follow-up compliance in WLHIV.

We anticipate a few limitations in the conduct of this study. The sample size for the *Phase 2* study may not be adequate to detect any significant difference in the risk of progression or persistence of CIN1 between all the screening methods being examined. The follow-up duration proposed in the study may also not be sufficient to allow a significant number of the enrolled women to progress from CIN1 to CIN2+. We, however, anticipate that despite these, the study will have significant public health impacts as this innovative method will offer a unique paradigm shift for future control of cervical cancer within an integrated HIV treatment setting in LMICs.

## Data Availability

No datasets were generated or analysed during the current study. All relevant data from this study will be made available upon study completion.

## Acknowledgements

The lead author (KSO) appreciates the mentorship support of Professor Ami S. Bhatt of Stanford University Medical School and Professor Folasade T. Ogunsola of the College of Medicine, University of Lagos. We also thank the staff of the Research Management Office (RMO) of the College of Medicine, University of Lagos for their assistance in securing the funding for this study.

## Statement of Ethics

This study protocol was reviewed and approved by the Health Research Ethics Committees of the Lagos University Teaching Hospital with approval number ADM/DSCST/HREC/APP/5204 and the College of Medicine, University of Lagos with approval number CMUL/HREC/07/22/1075. The research will be conducted ethically according to the World Medical Association Declaration of Helsinki. All potential study participants will give their written informed consent before enrolment and strict adherence to the privacy and confidentiality of participant’s information will be ensured during and after the conduct of the study.

## Supporting information caption

S1_Appendix – IRB approval (CMUL)

S2_Appendix – IRB approval (LUTH)

